# REACT-1 round 9 final report: Continued but slowing decline of prevalence of SARS-CoV-2 during national lockdown in England in February 2021

**DOI:** 10.1101/2021.03.03.21252856

**Authors:** Steven Riley, Haowei Wang, Oliver Eales, David Haw, Caroline E. Walters, Kylie E. C. Ainslie, Christina Atchison, Claudio Fronterre, Peter J. Diggle, Deborah Ashby, Christl A. Donnelly, Graham Cooke, Wendy Barclay, Helen Ward, Ara Darzi, Paul Elliott

## Abstract

**Background:** England will start to exit its third national lockdown in response to the COVID-19 pandemic on 8th March 2021, with safe effective vaccines being rolled out rapidly against a background of emerging transmissible and immunologically novel variants of SARS-CoV-2. A subsequent increase in community prevalence of infection could delay further relaxation of lockdown if vaccine uptake and efficacy are not sufficiently high to prevent increased pressure on healthcare services.

**Methods:** The PCR self-swab arm of the REal-time Assessment of Community Transmission Study (REACT-1) estimates community prevalence of SARS-CoV-2 infection in England based on random cross-sections of the population ages five and over. Here, we present results from the complete round 9 of REACT-1 comprising round 9a in which swabs were collected from 4th to 12th February 2021 and round 9b from 13th to 23rd February 2021. We also compare the results of REACT-1 round 9 to round 8, in which swabs were collected mainly from 6th January to 22nd January 2021.

**Results:** Out of 165,456 results for round 9 overall, 689 were positive. Overall weighted prevalence of infection in the community in England was 0.49% (0.44%, 0.55%), representing a fall of over two thirds from round 8. However the rate of decline of the epidemic has slowed from 15 (13, 17) days, estimated for the period from the end of round 8 to the start of round 9, to 31 days estimated using data from round 9 alone (lower confidence limit 17 days). When comparing round 9a to 9b there were apparent falls in four regions, no apparent change in one region and apparent rises in four regions, including London where there was a suggestion of sub-regional heterogeneity in growth and decline. Smoothed prevalence maps suggest large contiguous areas of growth and decline that do not align with administrative regions.

Prevalence fell by 50% or more across all age groups in round 9 compared to round 8, with prevalence (round 9) ranging from 0.21% in those aged 65 and over to 0.71% in those aged 13 to 17 years. Round 9 prevalence was highest among Pakistani participants at 2.1% compared to white participants at 0.45% and Black participants at 0.83%. There were higher adjusted odds of infection for healthcare and care home workers, for those working in public transport and those working in education, school, nursery or childcare and lower adjusted odds for those not required to work outside the home.

**Conclusions:** Community prevalence of swab-positivity has declined markedly between January and February 2021 during lockdown in England, but remains high; the rate of decline has slowed in the most recent period, with a suggestion of pockets of growth. Continued adherence to social distancing and public health measures is required so that infection rates fall to much lower levels. This will help to ensure that the benefits of the vaccination roll-out programme in England are fully realised.

## Introduction

Most European countries started 2021 with high numbers of COVID-19 cases [1] and stringent interventions [2]. During the first eight weeks of the year, cases fell across Europe before starting to increase again during February [1]. England entered its third national lockdown on 5th January 2021 [3] with the seven-day rolling average of cases at 95 per 100,000. By the 22nd February 2021, cases had fallen to an average of 18 per 100,000 during the previous seven days [4]. Over the same period, daily hospital admissions for COVID-19 fell from 3,592 to 949 (seven-day average) [4]. By the 22nd February, 15,113,158 people in England had received at least one dose [4] of either the BNT162b2 mRNA [5] or ChAdOx1 [6] COVID-19 vaccines.

On 22nd February 2021, the UK government announced a plan (roadmap) for the gradual easing of the lockdown to start on 8th March 2021 with the opening of schools before proceeding in four additional steps [7]. The roadmap leaves five-week minimum periods between steps to allow time for the impact of each set of relaxations on the epidemic to be assessed against four criteria: successful continuation of the vaccine roll-out programme, good efficacy of the vaccine against hospitalisations and deaths, no substantial change in the overall risk assessment of the pandemic because of SARS-CoV-2 variants, and no evidence that an increase in infections in the community may lead to a surge in hospitalisations such that healthcare services would be placed under unsustainable pressure.

Largely in line with cases, prevalence of infections in the community in England dropped substantially during January and early February 2021 [8,9]. Any large uptick in the prevalence of infections would signal a potential threat to the smooth continuation of the roadmap; but with high vaccine uptake [4] and encouraging early estimates of vaccine efficacy [5], the link between infections and strain on healthcare services will likely be weakened in the near future.

The PCR self-swab arm of the REal-time Assessment of Community Transmission Study (REACT-1) is designed to measure community prevalence of SARS-CoV-2 infection in England [10]. Here, we present results from the complete round 9 of REACT-1 comprising round 9a in which swabs were collected from 4th to 12th February 2021 and round 9b from 13th to 23rd February 2021. We also compare the results of REACT-1 round 9 to round 8, in which swabs were collected mainly from 6th January to 22nd January 2021.

## Results

REACT-1 round 9 included 165,456 individuals with a valid swab result of whom 689 tested positive, giving a weighted prevalence overall of 0.49% (0.44%, 0.55%), down by over two thirds from 1.57% (1.49%, 1.66%) in round 8 (Table 1). The round 9 data comprised 388 positives from 87,408 swabs in round 9a, with weighted prevalence of 0.51% (0.44%, 0.59%) and 301 positives from 78,047 swabs in round 9b, with a weighted prevalence slightly lower at 0.47% (0.40%, 0.55%).

**Table 1.**
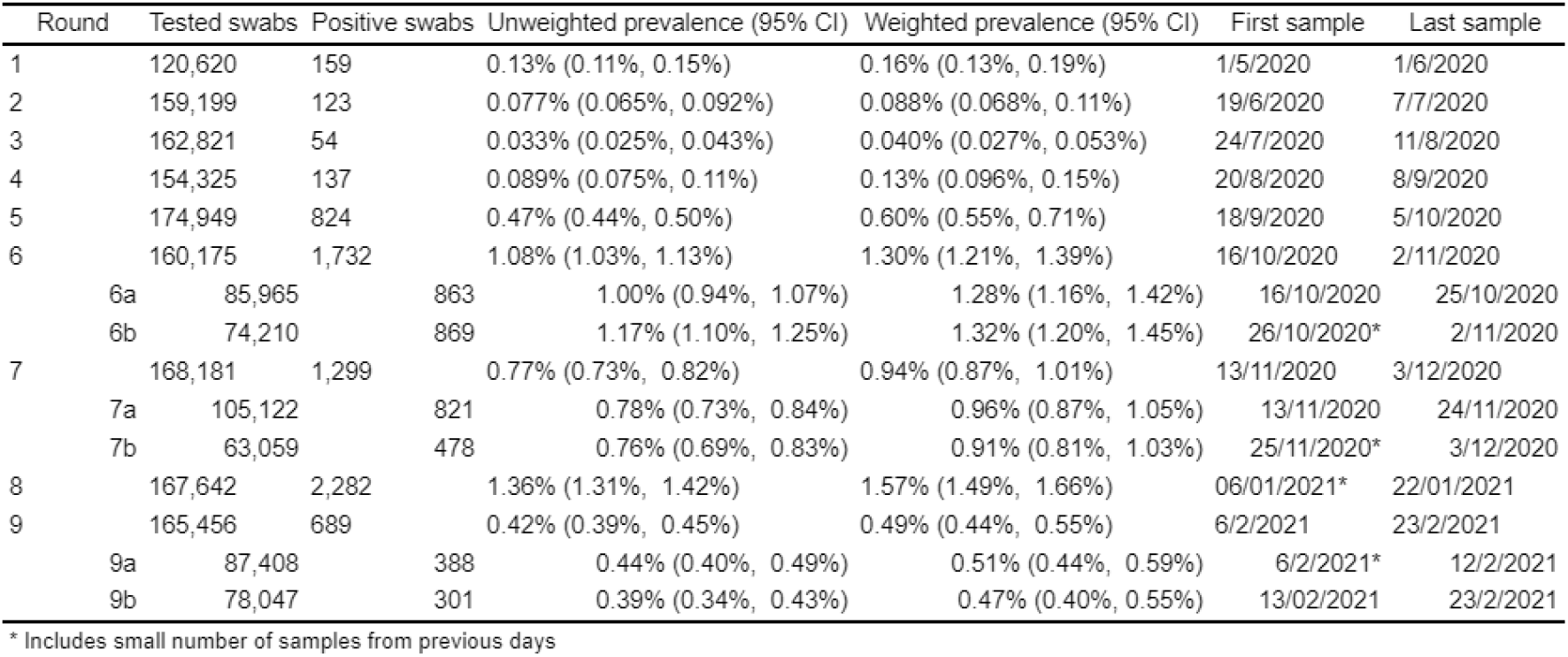
Unweighted and weighted prevalence of swab-positivity across nine rounds of REACT-1.

Using a constant growth rate model, we estimated halving times and R numbers for England using two time periods: from the second half of round 8 (8b) to round 9a and within round 9 (Table 2). We estimated a halving time of 15 (13, 17) days, corresponding to an R of 0.73 (0.69, 0.76) between rounds 8b and 9a, whereas from round 9a to 9b we estimated a halving time of 31 days (lower confidence limit 17 days) and a corresponding R of 0.86 (0.76, 0.97). We thus observe a slowing in the rate of decline of the epidemic in England on comparing these estimates (probability of difference in Rs > 0.99, not accounting for overlap in the time periods), although note that the estimate of R in the most recent period is still reliably below one. This slowing is reflected in the fitted P-spline which shows a flattening off in the most recent period (Figure 1).

**Table 2.** Estimates of national growth rates, doubling times and reproduction numbers for round 9a and 9b, and for round 8b and 9a.

| Round | Outcome | Growth rate | R | Probability R>1 | Growth(+)/Decay(-) rate |
| --- | --- | --- | --- | --- | --- |
| 9a and 9b | All positives | -0.022 ( -0.041 , -0.004 ) | 0.86 ( 0.76 , 0.97 ) | <0.01 | -30.9 ( -17.0 , * ) |
|  | Non-symptomatics | -0.028 ( -0.057 , 0.002 ) | 0.83 ( 0.67 , 1.01 ) | 0.03 | -25.2 ( -12.1 , * ) |
|  | Positive for both E and N genes | -0.036 ( -0.060 , -0.012 ) | 0.79 ( 0.66 , 0.93 ) | <0.01 | -19.5 ( -11.5 , * ) |
|  | Positive for both E and N genes or positive only for N gene with CT 35 or less | -0.022 ( -0.042 , -0.003 ) | 0.86 ( 0.75 , 0.98 ) | 0.01 | -30.8 ( -16.5 , * ) |
| 8b and 9a | All positives | -0.047 ( -0.054 , -0.040 ) | 0.73 ( 0.69 , 0.76 ) | <0.01 | -14.7 ( -12.8 , -17.2 ) |
|  | Non-symptomatics | -0.033 ( -0.045 , -0.021 ) | 0.80 ( 0.74 , 0.87 ) | <0.01 | -21.0 ( -15.5 , -33.3 ) |
|  | Positive for both E and N genes | -0.053 ( -0.062 , -0.044 ) | 0.69 ( 0.65 , 0.74 ) | <0.01 | -13.1 ( -11.2 , -15.6 ) |
|  | Positive for both E and N genes or positive only for N gene with CT 35 or less | -0.048 ( -0.055 , -0.040 ) | 0.72 ( 0.68 , 0.76 ) | <0.01 | -14.6 ( -12.6 , -17.3 ) |

\* Doubling/Halving time had an estimated magnitude greater than 50 days and so represented approximately constant prevalence

**Figure 1.**
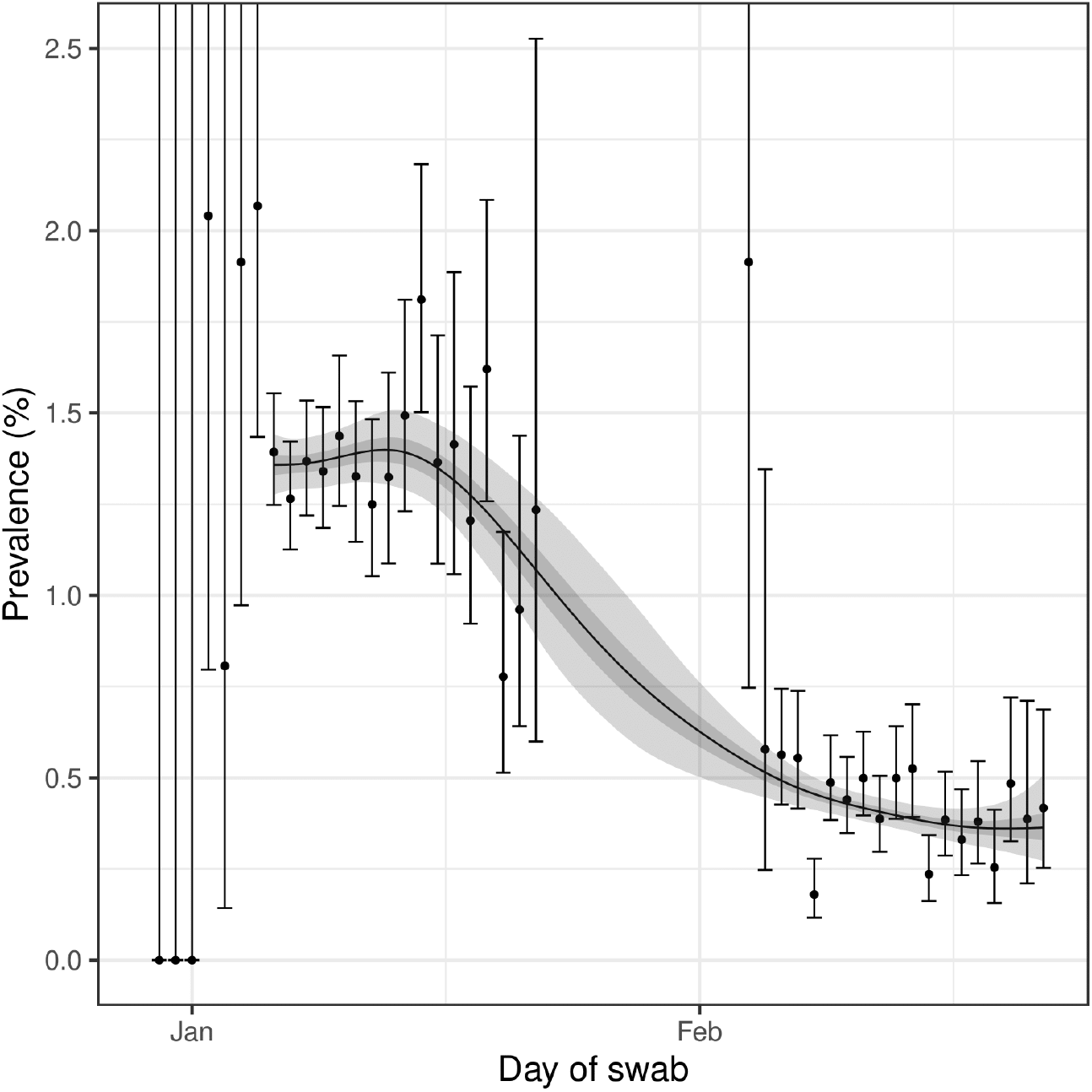
Prevalence of national swab-positivity for England estimated using a P-spline for all nine rounds with central 50% (dark grey) and 95% (light grey) posterior credible intervals. Shown here only for the period of round 8 to round 9. Unweighted observations (black dots) and 95% binomial confidence intervals (vertical lines) are also shown.

The decline in prevalence from round 8 to round 9 was seen in all age groups, which was 50% or more over this period (Table 3, Table 4, Figure 2). In the latter half of round 9 (9b), prevalence varied from 0.21% (0.14%, 0.31%) in those aged 65 and over to 0.71% (0.34%, 1.45%) in those aged 13 to 17 years.

**Table 3a.**
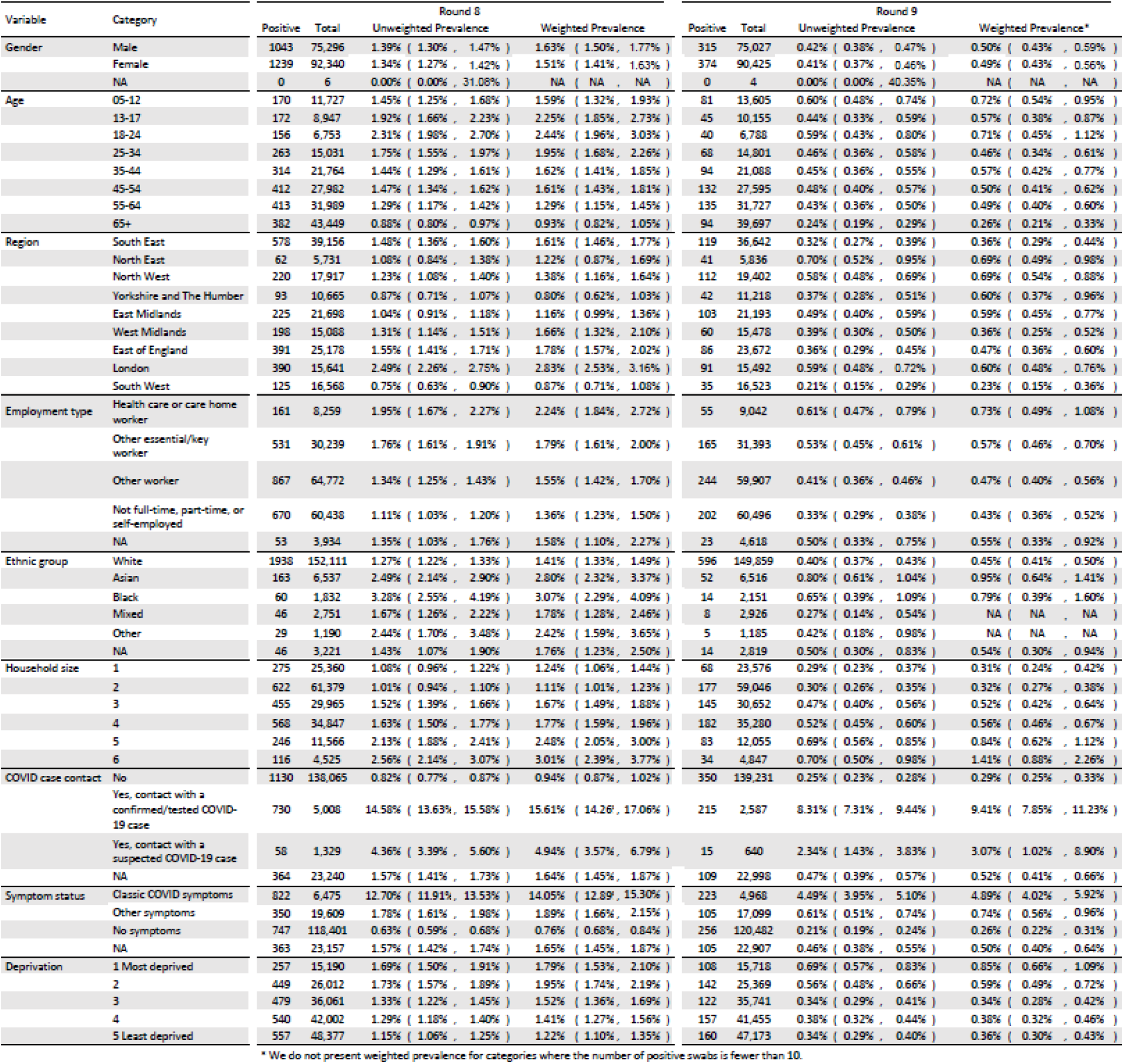
Unweighted and weighted prevalence of swab-positivity for core variables for rounds 8 and 9.

**Table 3b.**
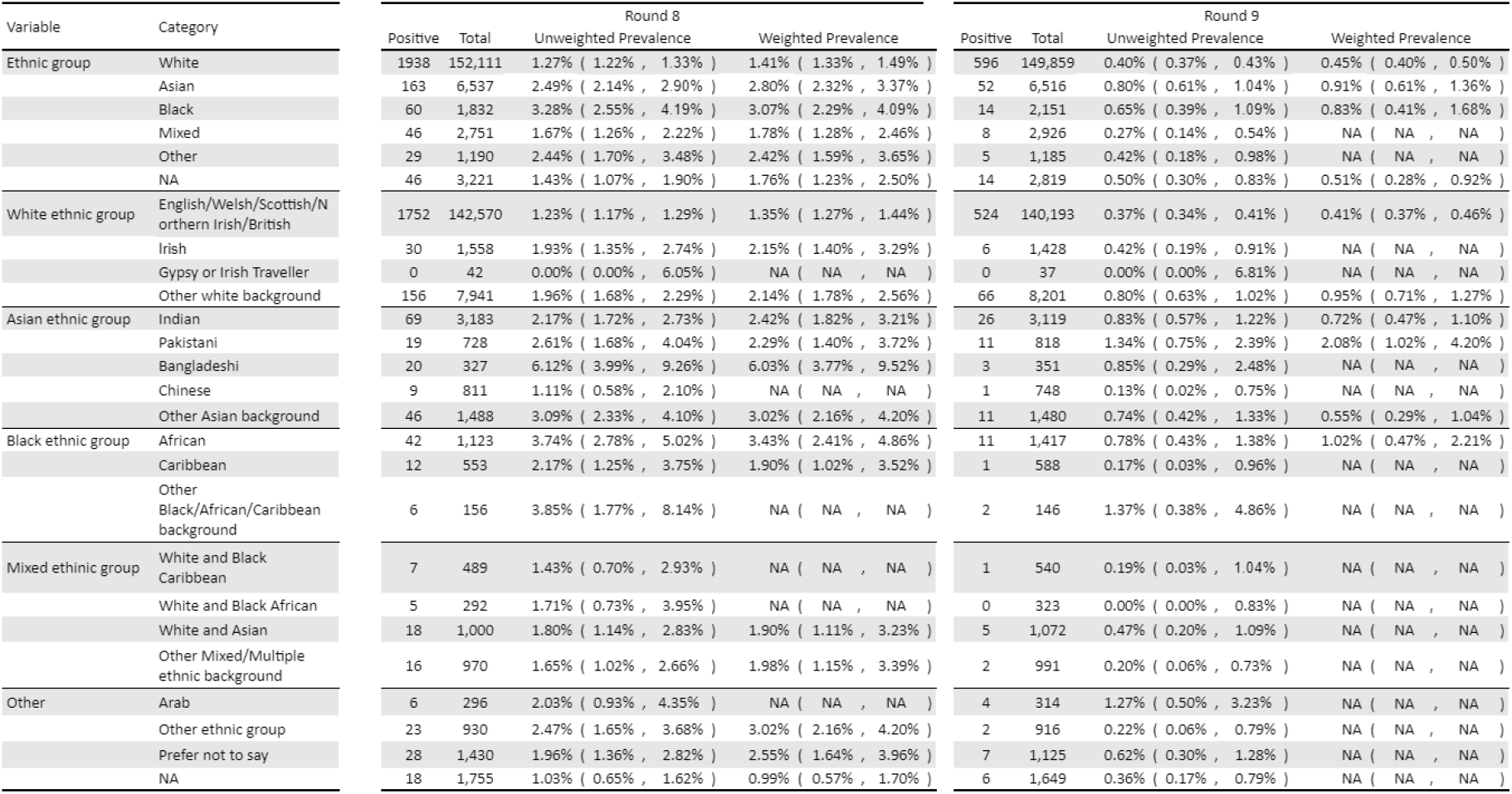
Unweighted and weighted prevalence of swab-positivity for detailed ethnicity categories for rounds 8 and 9.

**Table 3c.**
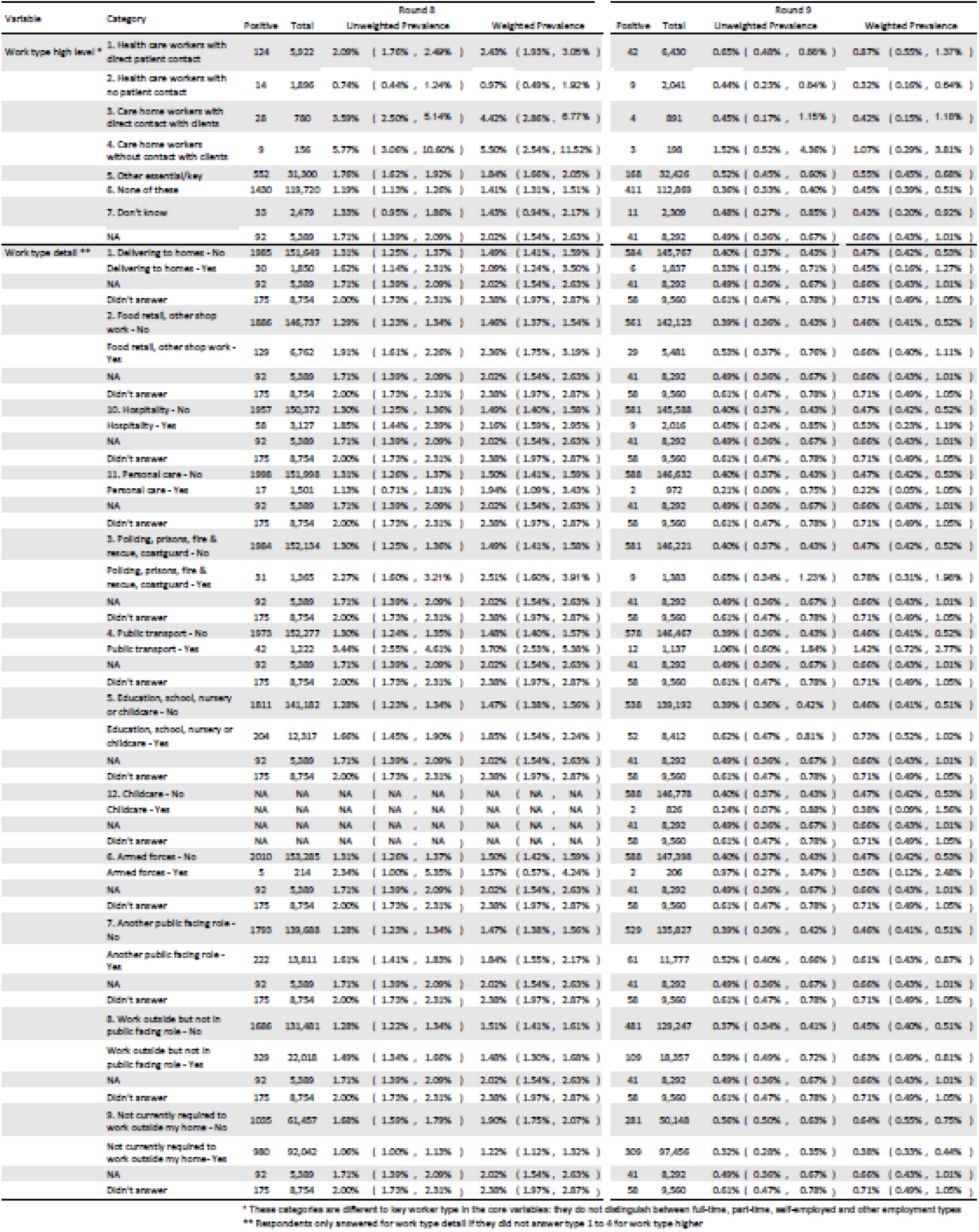
Unweighted and weighted prevalence of swab-positivity for detailed work types for respondents for rounds 8 and 9.

**Table 4.**
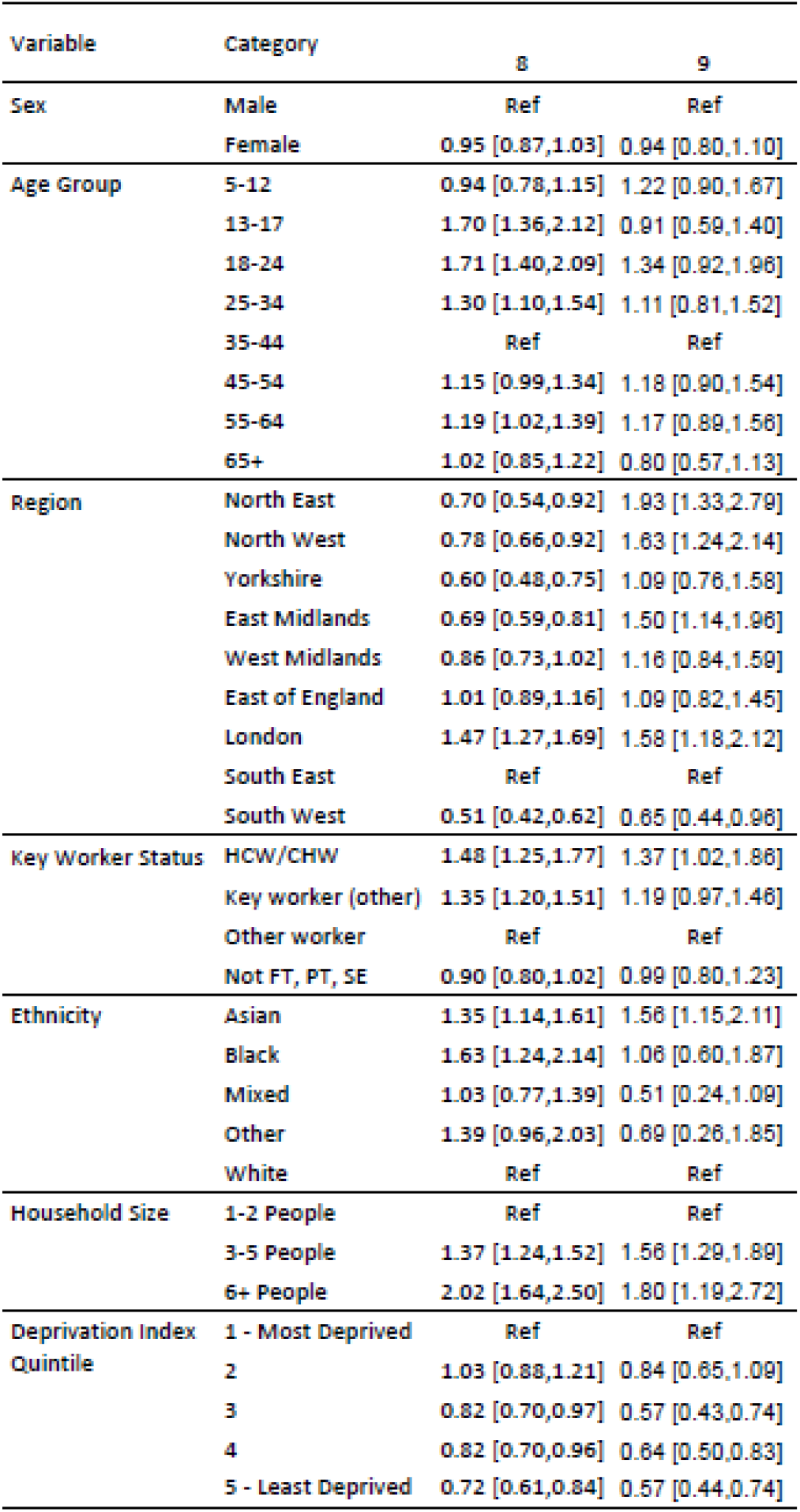
Mutually adjusted odds ratios for core variables for rounds 8 and 9. The deprivation index is based on the Index of Multiple Deprivation (2019) at lower super output area. Here we group scores into quintiles, where 1 = most deprived and 5 = least deprived. HCW/CHW = health care or care home workers; Not FT, PT, SE = Not full-time, part-time, or self-employed. Yorkshire = Yorkshire and The Humber.

**Figure 2.**
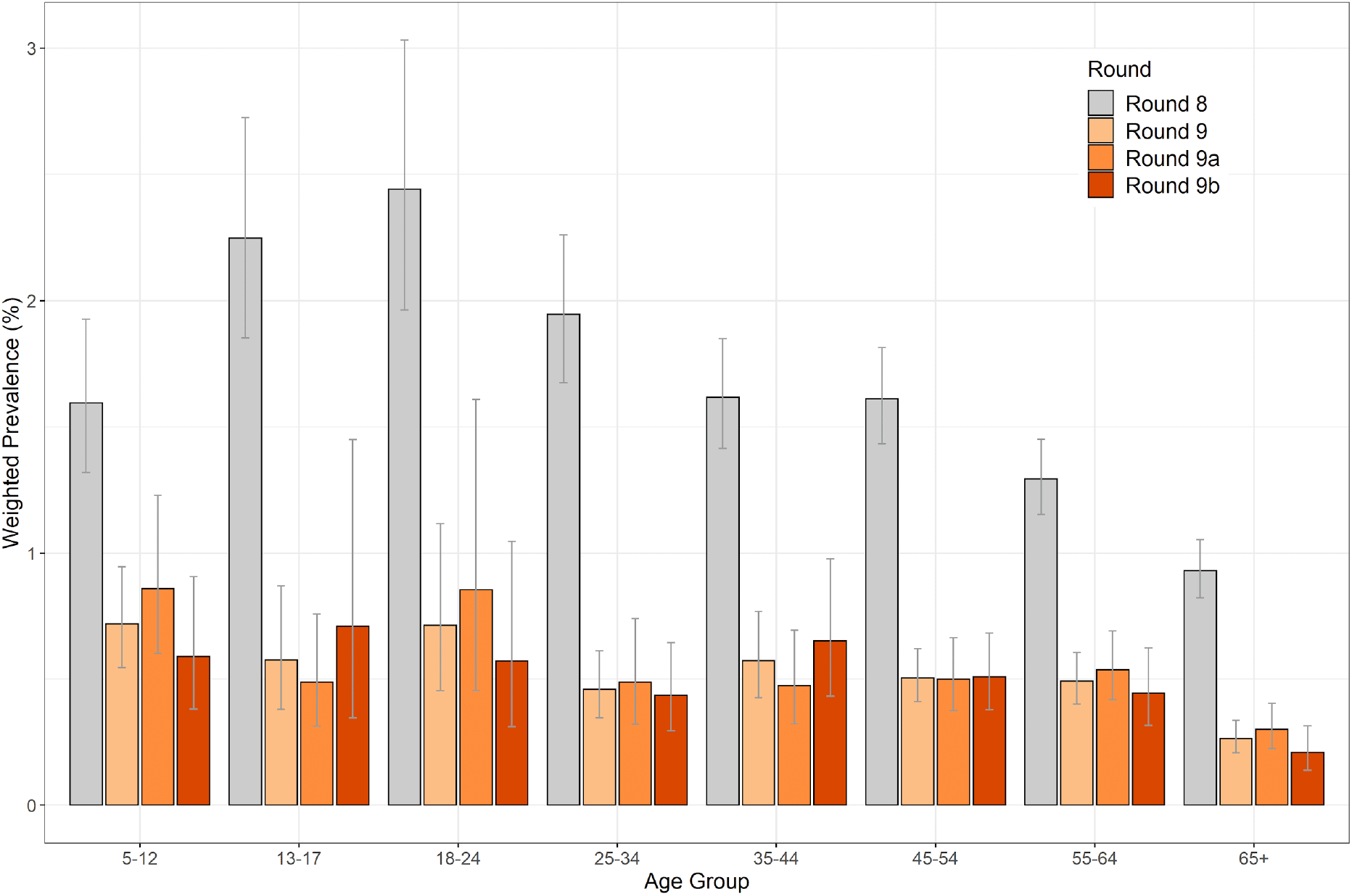
Weighted prevalence of swab-positivity by age groups for rounds 8, 9, 9a and 9b. Bars show 95% confidence intervals.

We observed differences in patterns of regional prevalence between rounds 8 and 9 when compared with patterns between rounds 9a and 9b (Table 3, Table 4, Figure 3). Between rounds 8 and 9 there were substantial falls in weighted prevalence in seven of the nine regions with smaller apparent falls in Yorkshire and The Humber and in North East. However, between rounds 9a and 9b, while there were apparent falls in North East, North West, East of England and South West, and no apparent change in Yorkshire and The Humber, there were apparent rises in London, South East, East Midlands and West Midlands. Using a constant regional growth rate model, we found evidence suggestive of growth (80% or greater probability) in London and South East and robust evidence of decline in North West (Table 5). These patterns were also reflected in the regional fitted P-splines (Figure 4).

**Table 5.**
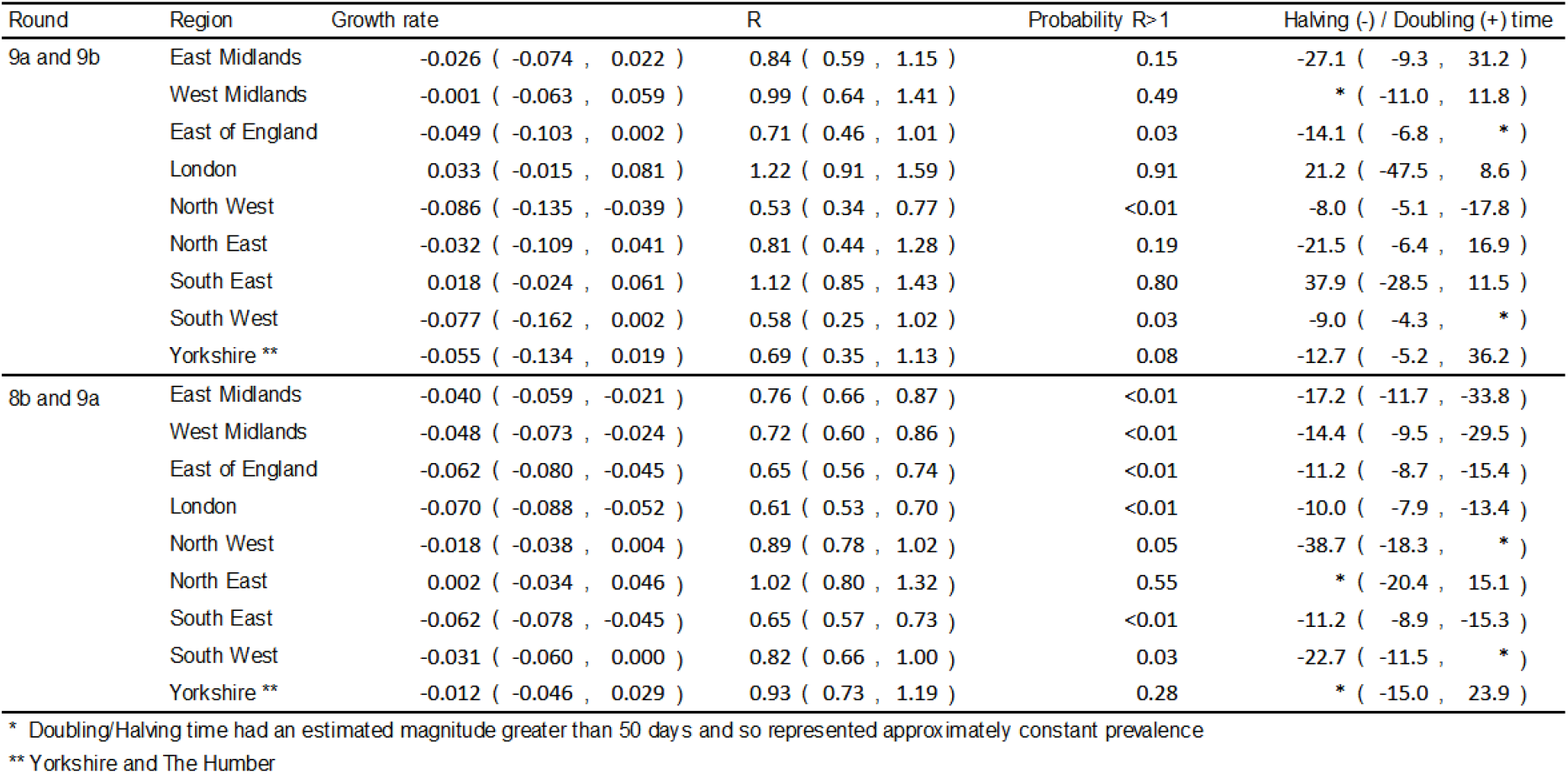
Estimates of regional growth rates, doubling times and reproduction numbers for round 9a and 9b, and for round 8b and 9a.

**Figure 3.**
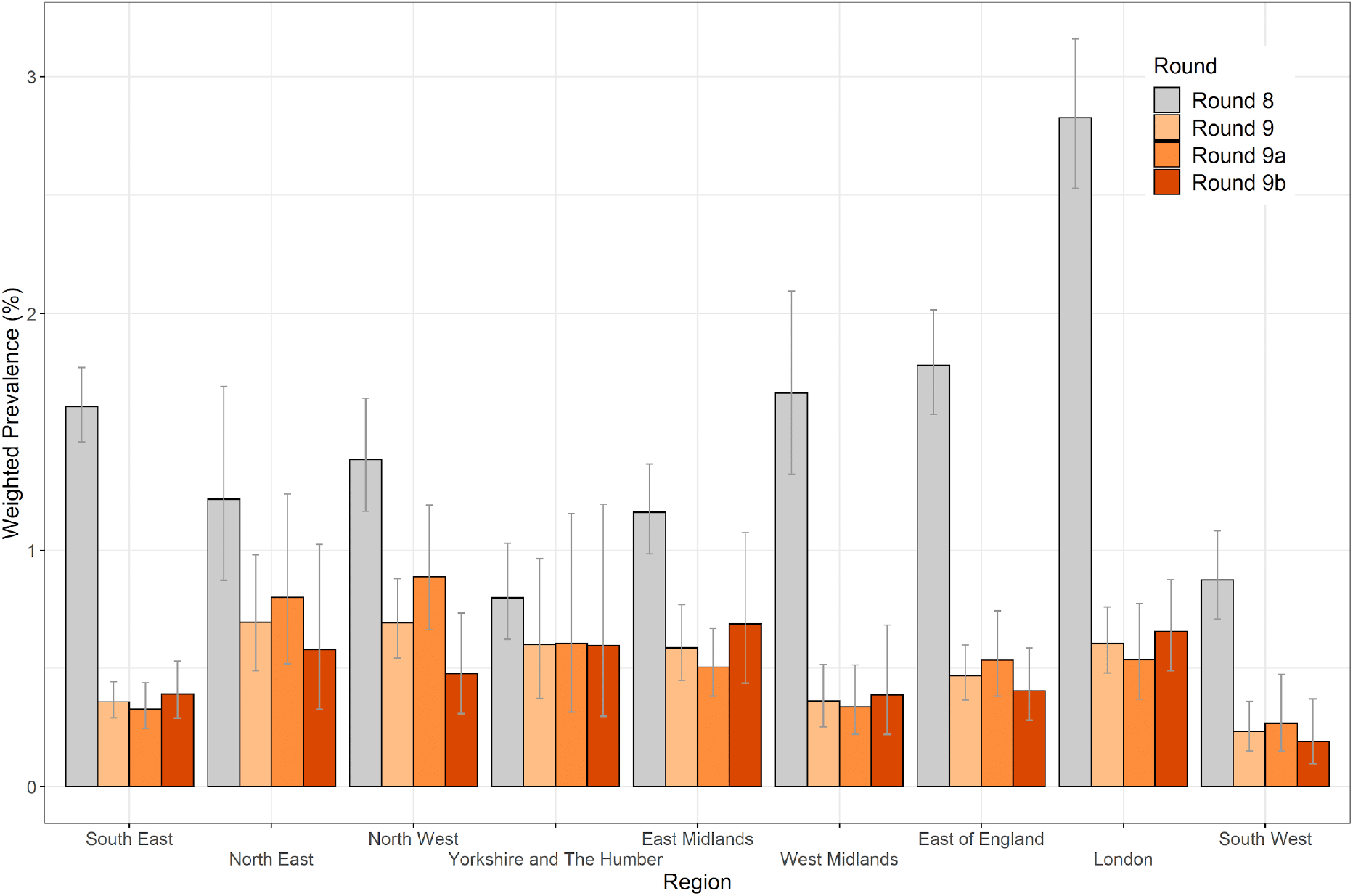
Weighted prevalence of swab-positivity by region for rounds 8, 9, 9a and 9b. Bars show 95% confidence intervals.

**Figure 4.**
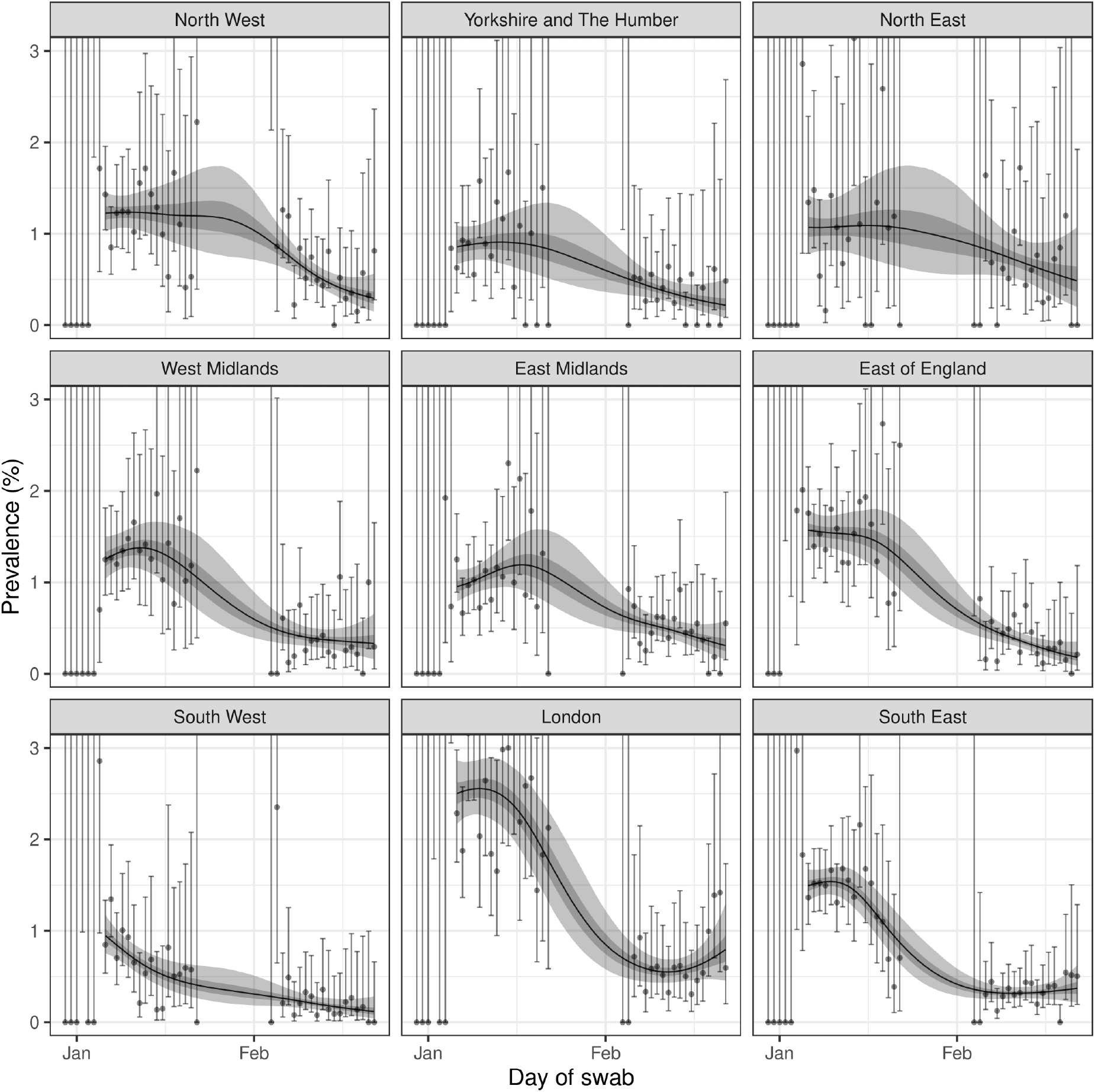
Prevalence of swab-positivity estimated using a P-spline (with constant second-order random walk prior) for each region of England separately. Each model was fit to all nine rounds but is only shown here for the period of round 8 to round 9. Central 50% (dark grey) and 95% (light grey) posterior credible intervals are also shown.

Maps for rounds 9a and 9b of unweighted, unsmoothed swab-positivity at the level of lower-tier local authorities (LTLA) suggest reductions in prevalence in eastern part of North West region, but are otherwise difficult to interpret with highly fragmented groups of high and low prevalence areas (Figure 5). We therefore used nearest neighbours within the study sample to smooth LTLA prevalence. We aimed to reveal underlying spatial structure and thus allow visualisation of apparent growth or decline. The resultant maps suggest large contiguous areas of apparent increasing or decreasing prevalence when comparing rounds 9a and 9b (Figure 6). A long tract of apparent increasing prevalence runs from the south coast, through south and west London (Figure 7) into the Midlands and then on to the west side of Yorkshire and the Humber. One contiguous area of decreasing prevalence was seen in north and east London, the southern part of East of England and the northern part of South East, with another in the western part of South West. We also observed sharp declines in the conurbations in the North West reflecting the overall decline in prevalence in that region.

**Figure 5.**
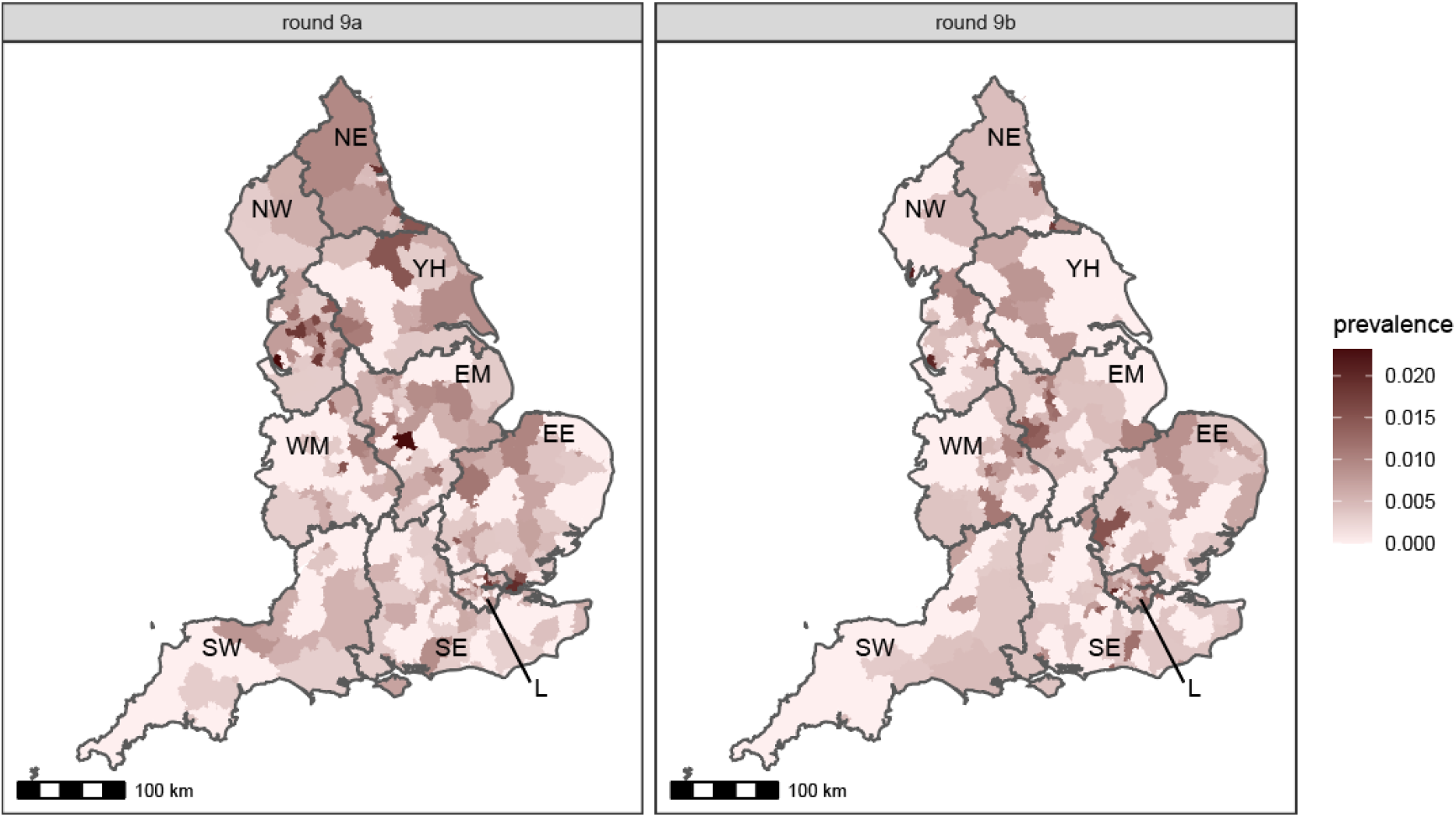
Unweighted, unsmoothed swab-positivity for lower-tier local authorities in England for rounds 9a and 9b of REACT-1. Regions: NE = North East, NW = North West, YH = Yorkshire and The Humber, EM = East Midlands, WM = West Midlands, EE = East of England, L = London, SE = South East, SW = South West.

**Figure 6.**
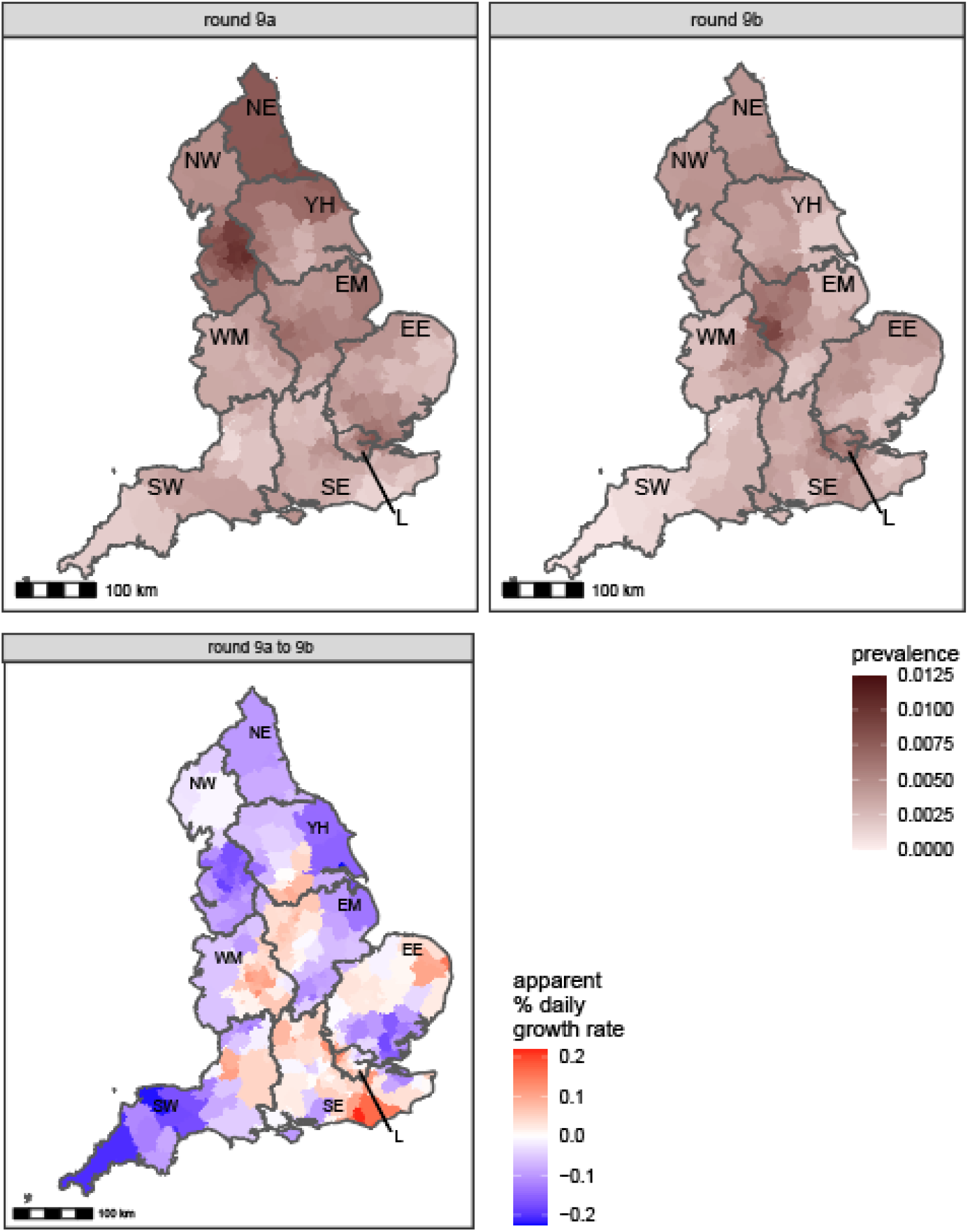
Neighbourhood prevalence of swab-positivity for rounds 9a and 9b (top), apparent percentage change in daily growth rate from round 9a to round 9b (bottom). Neighbourhood prevalence calculated from 2,532 (round 9a) and 2,193 (round 9b) nearest neighbours in the sample (the median number of neighbours within 30km in the study). Neighbourhood prevalence displayed as an average of a random sample of 15 participants for each lower-tier local authority (LTLA). Apparent percentage change in daily growth rate calculated as round 9b prevalence divided by round 9a prevalence divided by the difference in the median round date (in days). Regions: NE = North East, NW = North West, YH = Yorkshire and The Humber, EM = East Midlands, WM = West Midlands, EE = East of England, L = London, SE = South East, SW = South West.

**Figure 7.**
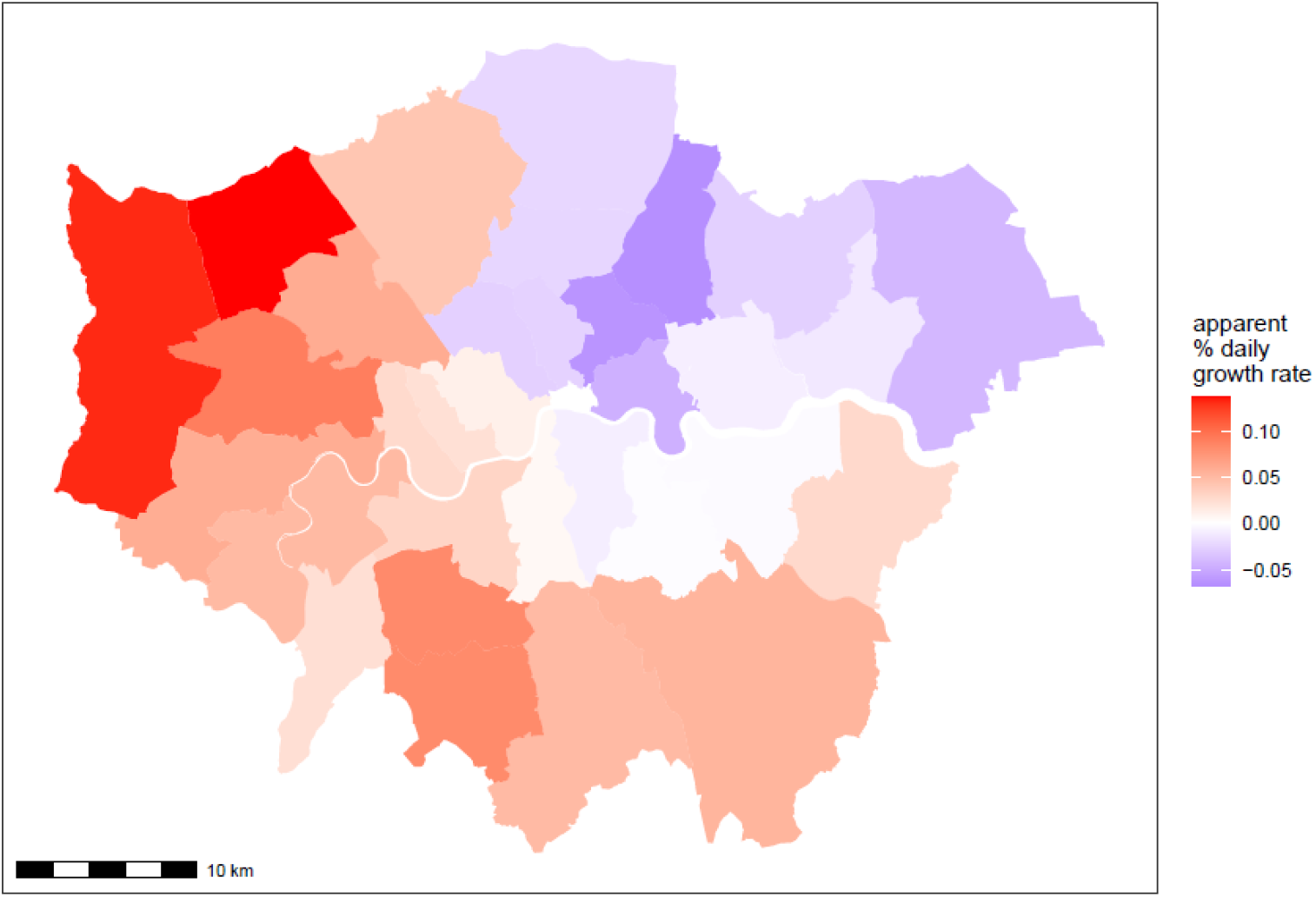
Apparent percentage change in daily growth rate from round 9a to round 9b for London, calculated as round 9b prevalence divided by round 9a prevalence divided by the difference in the median round date (in days).

We observed robust patterns in the prevalence of swab-positivity for ethnicity subgroups in both rounds 8 and 9 (Table 3b). In round 8, unweighted prevalence among Bangladeshi participants was very high at 6.1% (4.0%, 9.3%) compared to 1.2% (1.2%, 1.3%) in white participants. However unweighted prevalence in Bangladeshi fell to 0.85% (0.29%, 2.5%) in round 9 (we report unweighted prevalence for Bangladeshi participants because of small numbers of positives in round 9). In round 9, the highest weighted prevalence was amongst Pakistani participants at 2.1% (1.0%, 4.2%) compared with white participants at 0.41% (0.37%, 0.46%). Corresponding odds ratios adjusted for: age, sex, region, deprivation (core variables), showed similar patterns (Table 6).

**Table 6.** Mutually adjusted odds ratios among core variables and odds ratios for core-variable adjusted detailed ethnicity categories.

| Category | Round 8 | Round 9 |
| --- | --- | --- |
| English/Welsh/Scottish/Northern Irish/British | Ref | Ref |
| Irish | 1.41 [0.98,2.04] | 1.16 [0.52,2.61] |
| Gypsy or Irish Traveller | 0.00 [0.00,Inf] | 0.00 [0.00,Inf] |
| Other white background | 1.25 [1.05,1.49] | 2.01 [1.54,2.64] |
| White and Black Caribbean | 0.82 [0.39,1.74] | 0.39 [0.06,2.81] |
| White and Black African | 1.05 [0.43,2.55] | 0.00 [0.00,Inf] |
| White and Asian | 1.15 [0.72,1.84] | 1.08 [0.44,2.62] |
| Other Mixed/Multiple ethnic background | 1.01 [0.61,1.67] | 0.46 [0.11,1.86] |
| Indian | 1.36 [1.06,1.74] | 2.02 [1.35,3.02] |
| Pakistani | 1.58 [0.99,2.50] | 2.68 [1.46,4.93] |
| Bangladeshi | 3.03 [1.90,4.82] | 1.57 [0.50,4.96] |
| Chinese | 0.70 [0.36,1.36] | 0.33 [0.05,2.34] |
| Other Asian background | 1.81 [1.34,2.45] | 1.72 [0.94,3.16] |
| African | 1.92 [1.39,2.65] | 1.46 [0.79,2.70] |
| Caribbean | 1.24 [0.70,2.22] | 0.39 [0.05,2.76] |
| Other Black/African/Caribbean background | 2.25 [0.99,5.12] | 2.93 [0.72,11.94] |
| Arab | 1.18 [0.52,2.66] | 2.69 [0.99,7.28] |
| Other ethnic group | 1.51 [0.99,2.29] | 0.52 [0.13,2.09] |
| Prefer not to say | 1.36 [0.93,1.99] | 1.54 [0.72,3.25] |
| NA | 0.93 [0.58,1.49] | 1.08 [0.48,2.42] |

We also give prevalence and adjusted odds ratios for occupation for both rounds 8 and 9 (Table 3c, Table 4, Table 7). Healthcare workers and care home workers had higher adjusted odds of infection at 1.48 (1.25, 1.77) in round 8 and 1.37 (1.02, 1.86) in round 9 (Table 4) compared with other workers. Higher adjusted odds were seen in participants who worked in public transport at 2.17 (1.58, 2.97) in round 8, and 2.14 (1.20, 3.83) in round 9, compared with those who did not; higher adjusted odds were also seen in those working in education, school, nursery or childcare at 1.20 (1.03, 1.39) in round 8 and 1.43 (1.07, 1.91) in round 9 compared with participants not working in those settings (Table 7). Lower adjusted odds of swab-positivity were seen among those not currently required to work outside their home at 0.67 (0.61, 0.74) in round 8 and 0.64 (0.54, 0.76) in round 9 compared with those currently required to work outside their home.

**Table 7.** Odds ratios for detailed work types, unadjusted and adjusted for gender, age group, region, ethnicity and deprivation index.

| Variable | Category | Round 8 |  | Round 9 |  |
| --- | --- | --- | --- | --- | --- |
|  |  | Univariate | Adjusted | Univariate | Adjusted |
| Work type high level * | 1. Health care workers with direct patient contact | 1.19 [0.98,1.43] | 1.16 [0.95,1.41] | 1.26 [0.90,1.77] | 1.23 [0.87,1.73] |
|  | 2. Health care workers with no patient contact | 0.41 [0.24,0.71] | 0.42 [0.24,0.71] | 0.85 [0.43,1.67] | 0.89 [0.45,1.74] |
|  | 3. Care home workers with direct contact with clients | 2.07 [1.41,3.05] | 2.02 [1.37,2.98] | 0.87 [0.32,2.34] | 0.81 [0.30,2.18] |
|  | 4. Care home workers without contact with clients | 3.41 [1.73,6.72] | 3.52 [1.78,6.95] | 2.95 [0.94,9.33] | 3.04 [0.96,9.64] |
|  | 5. Other essential/key workers | Ref | Ref | Ref | Ref |
|  | 6. None of these | 0.67 [0.61,0.74] | 0.71 [0.64,0.78] | 0.70 [0.59,0.84] | 0.83 [0.69,1.00] |
|  | 7. Don't know | 0.75 [0.53,1.07] | 0.69 [0.48,0.99] | 0.92 [0.50,1.69] | 0.89 [0.48,1.64] |
|  | NA | 0.97 [0.77,1.21] | 0.53 [0.39,0.73] | 0.95 [0.68,1.34] | 0.93 [0.59,1.48] |
| Work type detail | 1. Delivering to homes - No | Ref | Ref | Ref | Ref |
|  | Delivering to homes - Yes | 1.24 [0.86,1.79] | 1.15 [0.80,1.63] | 0.81 [0.36,1.82] | 0.72 [0.32,1.61] |
|  | NA | 1.31 [1.06,1.62] | 0.75 [0.55,1.01] | 1.24 [0.90,1.70] | 1.08 [0.70,1.66] |
|  | Didn't answer | 1.54 [1.32,1.80] | 1.42 [1.21,1.67] | 1.52 [1.16,1.99] | 1.30 [0.99,1.71] |
|  | 2. Food retail, other shop work - No | Ref | Ref | Ref | Ref |
|  | Food retail, other shop work - Yes | 1.49 [1.25,1.79] | 1.30 [1.08,1.56] | 1.34 [0.92,1.95] | 1.13 [0.77,1.65] |
|  | NA | 1.33 [1.08,1.65] | 0.76 [0.56,1.04] | 1.25 [0.91,1.72] | 1.09 [0.70,1.68] |
|  | Didn't answer | 1.57 [1.34,1.83] | 1.44 [1.23,1.69] | 1.54 [1.17,2.02] | 1.31 [1.00,1.73] |
|  | 10. Hospitality - No | Ref | Ref | Ref | Ref |
|  | Hospitality - Yes | 1.43 [1.10,1.87] | 1.20 [0.92,1.56] | 1.12 [0.58,2.17] | 0.92 [0.47,1.78] |
|  | NA | 1.32 [1.07,1.63] | 0.76 [0.56,1.02] | 1.24 [0.90,1.70] | 1.08 [0.70,1.67] |
|  | Didn't answer | 1.55 [1.32,1.81] | 1.42 [1.21,1.67] | 1.52 [1.16,2.00] | 1.30 [0.99,1.72] |
|  | 11. Personal care - No | Ref | Ref | Ref | Ref |
|  | Personal care - Yes | 0.86 [0.53,1.39] | 0.81 [0.50,1.30] | 0.51 [0.13,2.06] | 0.47 [0.12,1.87] |
|  | NA | 1.30 [1.06,1.61] | 0.74 [0.55,1.01] | 1.23 [0.90,1.70] | 1.08 [0.70,1.66] |
|  | Didn't answer | 1.53 [1.31,1.79] | 1.41 [1.20,1.66] | 1.52 [1.16,1.99] | 1.30 [0.98,1.71] |
|  | 3. Policing, prisons, fire & rescue, coastguard - No | Ref | Ref | Ref | Ref |
|  | Policing, prisons, fire & rescue, coastguard - Yes | 1.76 [1.23,2.52] | 1.68 [1.17,2.41] | 1.64 [0.85,3.18] | 1.49 [0.77,2.90] |
|  | NA | 1.31 [1.06,1.62] | 0.75 [0.55,1.01] | 1.25 [0.91,1.71] | 1.08 [0.70,1.68] |
|  | Didn't answer | 1.54 [1.32,1.80] | 1.43 [1.22,1.68] | 1.53 [1.17,2.01] | 1.31 [1.00,1.73] |
|  | 4. Public transport - No | Ref | Ref | Ref | Ref |
|  | Public transport - Yes | 2.71 [1.99,3.70] | 2.17 [1.58,2.97] | 2.69 [1.52,4.78] | 2.14 [1.20,3.83] |
|  | NA | 1.32 [1.07,1.63] | 0.75 [0.56,1.02] | 1.25 [0.91,1.72] | 1.09 [0.70,1.68] |
|  | Didn't answer | 1.55 [1.33,1.82] | 1.43 [1.22,1.68] | 1.54 [1.18,2.02] | 1.32 [1.00,1.74] |
|  | 5. Education, school, nursery or childcare - No | Ref | Ref | Ref | Ref |
|  | Education, school, nursery or childcare - Yes | 1.30 [1.12,1.50] | 1.20 [1.03,1.39] | 1.60 [1.20,2.13] | 1.43 [1.07,1.91] |
|  | NA | 1.34 [1.08,1.65] | 0.77 [0.57,1.05] | 1.28 [0.93,1.76] | 1.12 [0.72,1.73] |
|  | Didn't answer | 1.57 [1.34,1.84] | 1.45 [1.23,1.70] | 1.57 [1.20,2.06] | 1.35 [1.02,1.79] |
|  | 12. Childcare - No | Ref | Ref | Ref | Ref |
|  | Childcare - Yes | NA | NA | 0.60 [0.15,2.42] | 0.57 [0.14,2.29] |
|  | NA | NA | NA | 1.24 [0.90,1.70] | 1.08 [0.70,1.67] |
|  | Didn't answer | NA | NA | 1.52 [1.16,1.99] | 1.30 [0.99,1.71] |
|  | 6. Armed forces - No | Ref | Ref | Ref | Ref |
|  | Armed forces - Yes | 1.80 [0.74,4.38] | 1.84 [0.76,4.48] | 2.45 [0.61,9.88] | 2.24 [0.55,9.07] |
|  | NA | 1.31 [1.06,1.61] | 0.75 [0.55,1.01] | 1.24 [0.90,1.70] | 1.08 [0.70,1.67] |
|  | Didn't answer | 1.54 [1.31,1.79] | 1.42 [1.21,1.67] | 1.52 [1.16,2.00] | 1.31 [0.99,1.72] |
|  | 7. Another public facing role - No | Ref | Ref | Ref | Ref |
|  | Another public facing role - Yes | 1.26 [1.09,1.45] | 1.21 [1.05,1.39] | 1.33 [1.02,1.74] | 1.22 [0.93,1.59] |
|  | NA | 1.34 [1.08,1.65] | 0.75 [0.56,1.02] | 1.27 [0.92,1.75] | 1.09 [0.71,1.69] |
|  | Didn't answer | 1.57 [1.34,1.84] | 1.45 [1.23,1.70] | 1.56 [1.19,2.05] | 1.33 [1.01,1.76] |
|  | 8. Work outside but not in public facing role - No | Ref | Ref | Ref | Ref |
|  | Work outside but not in public facing role - Yes | 1.17 [1.04,1.32] | 1.12 [0.99,1.27] | 1.60 [1.30,1.97] | 1.48 [1.19,1.83] |
|  | NA | 1.34 [1.08,1.65] | 0.75 [0.56,1.01] | 1.33 [0.97,1.83] | 1.12 [0.72,1.73] |
|  | Didn't answer | 1.57 [1.34,1.84] | 1.44 [1.23,1.69] | 1.63 [1.24,2.15] | 1.39 [1.05,1.83] |
|  | 9. Not currently required to work outside my home - No | Ref | Ref | Ref | Ref |
|  | Not currently required to work outside my home- Yes | 0.63 [0.58,0.69] | 0.67 [0.61,0.74] | 0.56 [0.48,0.66] | 0.64 [0.54,0.76] |
|  | NA | 1.01 [0.82,1.26] | 0.60 [0.44,0.82] | 0.88 [0.63,1.22] | 0.80 [0.51,1.26] |
|  | Didn't answer | 1.19 [1.01,1.40] | 1.17 [0.99,1.38] | 1.08 [0.82,1.44] | 1.03 [0.77,1.37] |

\* These categories do not distinguish between full-time, part-time, self-employed and other employment types, as is the case in the core variable worker type categories.

Community prevalence of infection, as measured by REACT-1, shows strong apparent correlation with hospital admissions in England. With a fitted lag of 18 (18, 20) days between dates of swab and admissions, the national trend in swab-positivity and hospital admissions are well-aligned (Figure 8). Best-fit lags were slightly different across the age groups: 19 (19, 19) days for 6-17 year olds, 22 (22, 22) days for 18-64 year olds, 20 (18, 21) days for 65-84 year olds and 16 (12, 29) for 85+ year olds.

**Figure 8.**
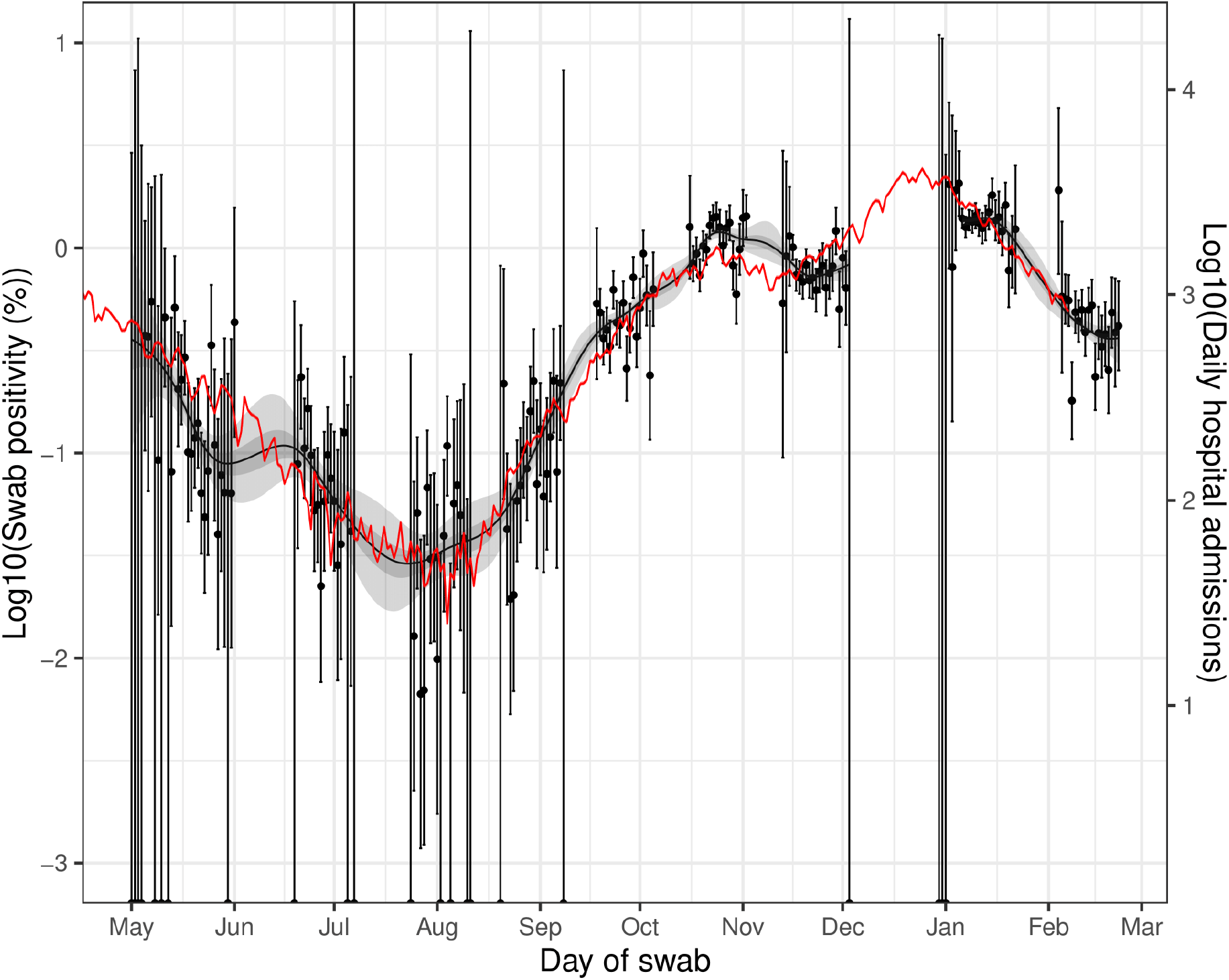
Daily hospital admissions in England (Solid red line, right hand y-axis) shifted by a lag parameter along the x-axis (see below), and daily swab positivity for all 9 rounds of the study (dots with 95% confidence intervals, left hand y-axis) and the P-spline estimate for swab positivity (Solid black line, dark grey shaded area is 50% central credible interval, light grey shaded area is 95% central credible interval, left-hand y-axis). Daily hospital admissions have been fit to observations from the REACT-1 study to obtain scaling and lag parameters. These parameter values were estimated using a Bayesian MCMC model: daily_positives(t) ∼ Binomial(daily_swab_tests(t), p = daily_admissions(t+lag)*scale). The time lag parameter was estimated at 18 (18, 20) days. Note the P-spline is not plotted for the region between round 7 and 8 in which there was an unobserved peak in swab-positivity.

**Figure 9.**
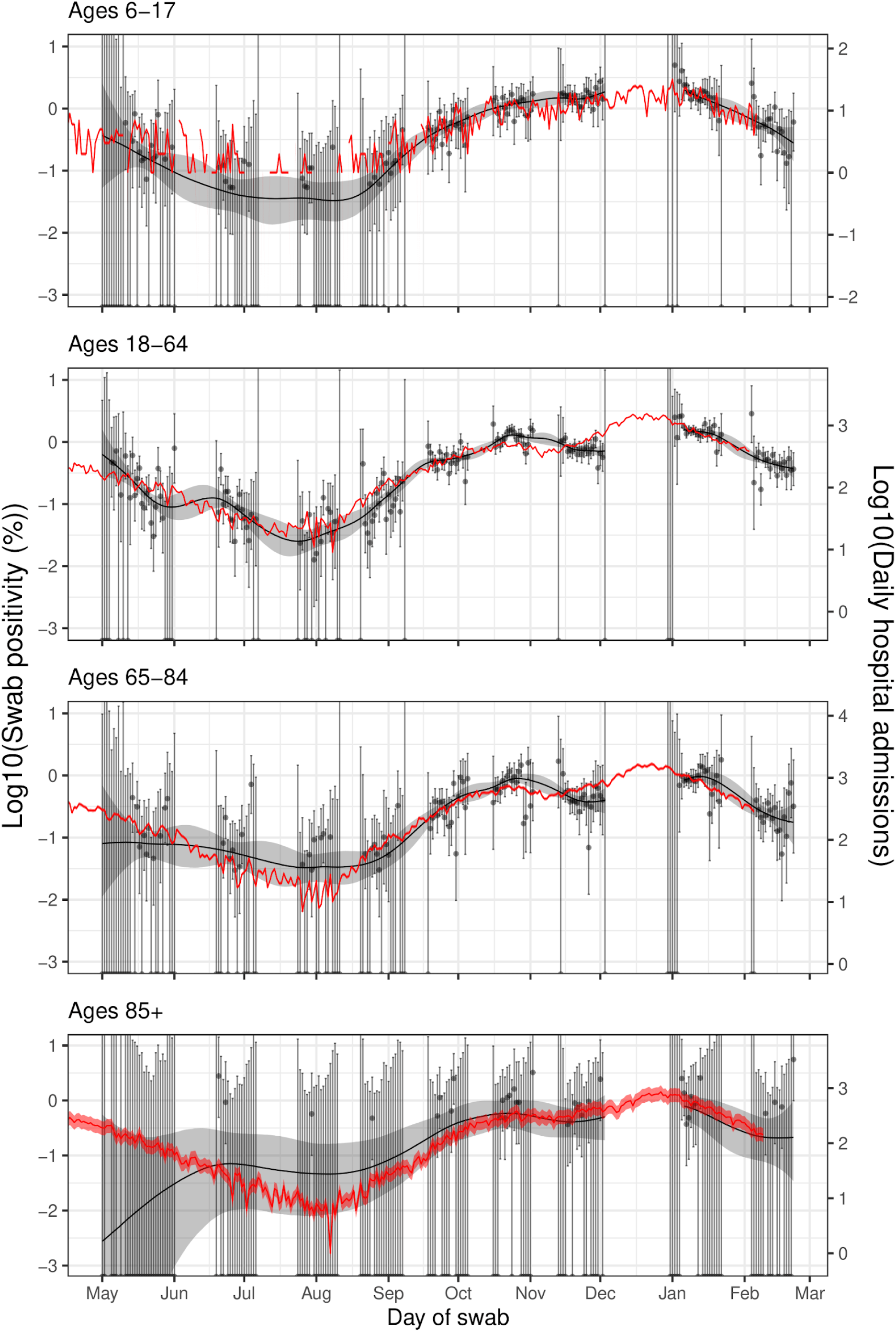
Daily hospital admissions in England for age groups 6-17, 18-64, 65-84 and 85+(Solid red line, right hand y-axis) shifted by 19, 22, 20 and 16 days along the x-axis respectively. Also plotted is the daily swab positivity for all 9 rounds of the study (Points with 95% confidence intervals, left hand y-axis) and the P-spline estimate for swab positivity (Solid black line, light grey shaded area is 95% central credible interval, left-hand y-axis). Note the p-spline is not plotted for the region between round 7 and 8 in which there was a peak that we do not have data to capture. Daily hospital admissions have been multiplied by a scaling parameter, alpha, and shifted by a time lag parameter, tau in order to correspond to the left hand y-axis. These parameter values were estimated using an MCMC with likelihood function: daily_swab_positives(t) ∼ Binomial(daily_swab_tests(t), p = daily_hospital_admissions(t+tau)*alpha). Estimated time lag parameters were 19 (19, 19) days for 6-17 year olds, 22 (22, 22) days for 18-64 year olds, 20 (18, 21) days for 65-84 year olds and 16 (12, 29) for 85+ year olds. Shaded red region shows the uncertainty in the parameter alpha and by how much daily hospital admissions should be scaled to correspond to swab positivity on the left hand y-axis. Note that for 6-17 year olds some days had zero hospital admissions and so these have not been included on the log scale graph.

## Discussion

Round 9 of the REACT-1 study was carried out during February 2021, some four to seven weeks after the beginning of the third national lockdown in England. We observed a marked decline in prevalence compared to the high levels seen at the beginning of the lockdown (round 8) [8], although the rate of decline slowed nationally during the most recent period. These results are consistent with community testing data (Pillar 2) which have likewise shown a slowing in the decline in prevalence over the recent period [4].

At regional scale, we observed a suggestion of plateauing or small rise in some areas, most notably in London, where there appeared to be sub-regional heterogeneity in the rates of growth and decline – while prevalence in north and east London appeared to be declining, there were apparent increases in parts of west and south London. Prevalence also appeared to be increasing in parts of West and East Midlands and the east coast of England, while there were continued sharp declines in North West conurbations which had seen high levels of infection during the second wave. Continued vigilance is necessary to ensure that the evident gains achieved in lockdown are maintained as lockdown measures are eased over coming weeks.

Unlike our previous reports, we show here results for more detailed categories of ethnicity and occupation. The highest prevalence by ethnicity across rounds 8 and 9 was found among Bangladeshi participants in whom unweighted prevalence reached 6% in January, before falling to less than one percent in February 2021. There were also high rates among Black, Indian and Pakistani compared to white participants in January 2021 when infections were still near their peak in the second wave. We found modestly higher adjusted odds of infection among those working in healthcare and care home settings and in education, school, nursery or childcare, whereas those working in public transport had over 2-fold increased adjusted odds of infection. In contrast, we found adjusted odds lower by approximately one third in those who were not required to work outside the home, stressing the importance of working from home during lockdown where possible to minimise social contacts and hence reduce the risk of transmission.

Our findings in February 2021 are possibly too early to detect the effect of the vaccination programme on rates of infection. The vaccination programme began to be rolled out in earnest from late December 2020 and early January 2021 in England, and to date over 17 million adults in England have received their first dose of vaccine [4]. Results from population studies indicate that one dose of either BNT162b2 mRNA or ChAdOx1 COVID-19 vaccines confers not only a reduction in risk of severe infection and hospitalisation but is also protective against symptomatic infection [11].

We show that trends in hospitalisations for COVID-19 closely match those of community SARS-CoV-2 infection prevalence in REACT-1 but with a lag period of around 16 to 22 days depending on age. We may expect to see a divergence in these curves in future rounds to the extent that there is an uncoupling between risk of infection and risk of severe disease and hospitalisation as a result of the vaccination programme. Specifically, we note that if vaccination has higher age-specific efficacy against hospital admission than efficacy against being found swab positive, we would expect the correlation between infection prevalence and hospital admissions to decrease in the affected age groups. On the other hand, should new variants subsequently emerge against which vaccines are less effective, or should protection from natural or vaccine-induced immunity wane, we would expect the correlation between community infection prevalence and hospital admissions to increase.

Our study has limitations. We include randomly selected cross-sections of the population of England and as such our estimates include people without symptoms as well as symptomatic people who are eligible for routine testing. We are therefore able to track the spread of the virus at population level and not rely on presentation of individuals to the routine testing programme. Our sampling method is designed to provide reliable estimates of prevalence at the small-area (LTLA) scale; however this does mean that we over-sample more rural areas and under-sample more urban areas, requiring re-weighting to obtain prevalence estimates that are representative of the country as a whole. As the vaccination programme is extended across the adult population, there may in the future be a greater reluctance to take part in our study if people feel they are protected from infection based on vaccination history. In addition, schools-based testing using lateral flow tests is being implemented in England which may again have an impact on participation rates. However, to date engagement with the programme has been high with swabs obtained from 22% to over 30% of people invited by letter to join the study.

In conclusion, community prevalence of swab-positivity has declined markedly between January and February 2021 during lockdown in England, but remains high; the rate of decline has slowed in the most recent period, with a suggestion of pockets of growth. Continued adherence to social distancing and public health measures is required so that infection rates fall to much lower levels. This will help to ensure that the benefits of the vaccination roll-out programme in England are fully realised.

## Methods

In REACT-1, we invite a random sample of the population in England (ages 5 years and over) to undertake a self-administered throat and nose swab (parent/guardian assisted at ages 5 to 12 years) and complete a questionnaire on demographics, health and lifestyle [10]. Swabs are picked up by courier and sent chilled to a single laboratory for RT-PCR. We use the National Health Service patient register to obtain the sample, aiming to obtain approximately equal numbers of participants in each of the 315 LTLAs in England.

In round 9, we sent out 761,000 letters to named individuals, with 210,046 (27.6%) kits dispatched. From these, we received completed swabs with a valid test result from 165,458 (78.8%) individuals giving an overall response rate (valid swabs divided by total number of people invited) of 21.7%.

We calculate prevalence of RT-PCR swab-positivity both unweighted and weighted to take account of the sample design and variable non-response, aiming to be representative of the population of England as a whole, by age, sex, region and ethnicity. We estimate prevalence by region, socio-demographic, occupational and other characteristics, as well as odds of swab-positivity based on a multivariable logistic regression model to account for potential confounding.

We use exponential growth models to estimate the reproduction number R, across and within rounds, both at national and regional levels. We examine trends over time by plotting the daily prevalence of swab-positivity and fitting a smoothed P-spline function to these daily prevalence data with knots at 5-day intervals [12]. We map the smoothed geographic variation in prevalence at LTLA level by use of a neighbourhood spatial smoothing method (scale: up to 30 km) across nearest neighbours as described in Figure 6.

We include sensitivity analyses in the estimation of R using different cut-points of cycle threshold (CT) values for determining swab-positivity, and by consideration only of individuals who did not report symptoms in the previous week. We also compare our data over time with those on hospitalisations from the Office for National Statistics.

We carried out the statistical analyses in R [13]. We obtained research ethics approval from the South Central-Berkshire B Research Ethics Committee (IRAS ID: 283787).

## Data Availability

The datasets generated or analysed, or both, during this study are not publicly available because of governance restrictions.

## Data availability

Supporting data for tables and figures are available either: in this Google [spreadsheet](https://docs.google.com/spreadsheets/d/1FsEnOKc_IUmOaEkIXzyuqr6jy9_3zLNI00wQuXNkgUA/edit?usp=sharing); or in the inst/extdata directory of this [GitHub R package](https://github.com/mrc-ide/reactidd).

## Acknowledgements

SR, CAD acknowledge support: MRC Centre for Global Infectious Disease Analysis, National Institute for Health Research (NIHR) Health Protection Research Unit (HPRU), Wellcome Trust (200861/Z/16/Z, 200187/Z/15/Z), and Centres for Disease Control and Prevention (US, U01CK0005-01-02). GC is supported by an NIHR Professorship. HW acknowledges support from an NIHR Senior Investigator Award and the Wellcome Trust (205456/Z/16/Z). PE is Director of the MRC Centre for Environment and Health (MR/L01341X/1, MR/S019669/1). PE acknowledges support from Health Data Research UK (HDR UK); the NIHR Imperial Biomedical Research Centre; NIHR HPRUs in Chemical and Radiation Threats and Hazards, and Environmental Exposures and Health; the British Heart Foundation Centre for Research Excellence at Imperial College London (RE/18/4/34215); and the UK Dementia Research Institute at Imperial (MC_PC_17114). We thank The Huo Family Foundation for their support of our work on COVID-19.

We thank key collaborators on this work – Ipsos MORI: Kelly Beaver, Sam Clemens, Gary Welch, Nicholas Gilby, Kelly Ward and Kevin Pickering; Institute of Global Health Innovation at Imperial College: Gianluca Fontana, Sutha Satkunarajah, Didi Thompson and Lenny Naar; Molecular Diagnostic Unit, Imperial College London: Prof. Graham Taylor; North West London Pathology and Public Health England for help in calibration of the laboratory analyses; Patient Experience Research Centre at Imperial College and the REACT Public Advisory Panel; NHS Digital for access to the NHS register; and the Department of Health and Social Care for logistic support. SR acknowledges helpful discussion with attendees of meetings of the UK Government Office for Science (GO-Science) Scientific Pandemic Influenza – Modelling (SPI-M) committee.

